# Analyzing human knockouts to validate GPR151 as a therapeutic target for reduction of body mass index

**DOI:** 10.1101/2021.10.21.21264378

**Authors:** Allan Gurtan, John Dominy, Shareef Khalid, Linh Vong, Shari Caplan, Treeve Currie, Sean Richards, Lindsey Lamarche, Daniel Denning, Diana Shpektor, Anastasia Gurinovich, Asif Rasheed, Shahid Hameed, Subhan Saeed, Imran Saleem, Anjum Jalal, Shahid Abbas, Raffat Sultana, Syed Zahed Rasheed, Fazal-ur-Rehman Memon, Nabi Shah, Mohammad Ishaq, Amit Khera, John Danesh, Sekar Kathiresan, Philippe Frossard, Danish Saleheen

## Abstract

Novel drug targets for sustained reduction in body mass index (BMI) are needed to curb the epidemic of obesity, which affects 650 million individuals worldwide and is a causal driver of cardiovascular and metabolic disease and mortality. Previous studies reported that the Arg95Ter nonsense variant of GPR151, an orphan G protein-coupled receptor, is associated with reduced BMI and reduced risk of Type 2 Diabetes (T2D). Here, we follow up on *GPR151* with the Pakistan Genome Resource (PGR), which is one of the largest exome biobanks of human homozygous loss-of-function carriers (knockouts) in the world. Among PGR participants, we identify 3 GPR151 putative loss-of-function (plof) variants (Arg95Ter, Tyr99Ter, and Phe175LeufsTer7) with a cumulative allele frequency of 2.2% and present at homozygosity. We confirm these alleles *in vitro* as loss-of-function. We test if *GPR151* plof is associated with BMI, T2D, or other metabolic traits. *GPR151* deficiency in complete human knockouts is not associated with a clinically significant difference in BMI. Moreover, loss of *GPR151* confers a nominally significant increase in risk of T2D (odds ratio = 1.2, p value = 0.03). Relative to wild-type mice, *Gpr151*^*-/-*^ animals exhibit no difference in body weight on normal chow, and higher body weight on a high-fat diet, consistent with the findings in humans. Together, our findings indicate that GPR151 antagonism is not a compelling therapeutic approach for obesity.

## Introduction

Obesity, defined as a body mass index (BMI) of >30 kg/m^2^, is a major global health concern. In 2015, 7.1% of global deaths were attributable to high BMI (Collaborators et al. 2017). Predictions estimate that half of the world’s population will be obese by 2030 (Tremmel et al. 2017). By 2030 in the United States alone, 25% of the population may be severely obese, as defined by BMI>35 kg/m^2^ (Ward et al. 2019). Being overweight or obese leads to a steep increase in all-cause mortality (Global et al. 2016), largely through increasing risk for Type 2 Diabetes (T2D) (Al-Salameh et al. 2019), nonalcoholic fatty liver disease (NAFLD) (Diehl and Day 2018), and cardiovascular disease (CVD) (Collaborators et al. 2017). Reduction in BMI may promote remission of multiple conditions (Wolfe et al. 2016).

Given the mortality, morbidity and public cost associated with obesity, there is a strong interest in identifying drug targets for sustained reduction in BMI. To date, two particular therapies that promote body weight reduction also reduce hospitalization and mortality from associated co-morbidities. Glucagon-like peptide-1 (GLP-1) agonists are incretin mimetics that can reduce body weight by up to 12% (Wilding et al. 2021), improve glycemic status (Niman et al. 2021), and reduce cardiovascular events (Niman et al. 2021). SGLT2 inhibitors prevent glucose re-uptake in the kidneys, reduce body weight by 2 kg, improve glycemic status, reduce cardiovascular events, and improve kidney function (Neal et al. 2017; McMurray et al. 2019; Wiviott et al. 2019; Cannon et al. 2020; Heerspink et al. 2020; Packer et al. 2020). Both GLP-1 agonists and SGLT2 inhibitors modify glucose metabolism, nonetheless, their effect on body weight is consistent with the expectation that reduction of BMI is therapeutically beneficial.

Human studies have identified numerous genetic loci associated with BMI (Locke et al. 2015). However, many of the genes linked to these loci are either technically challenging to drug or are poorly validated as causal. For example, the fat mass and obesity-associated (FTO) gene encodes an mRNA demethylase (Gerken et al. 2007; Jia et al. 2011) strongly associated with BMI (Frayling et al. 2007). However a direct role for FTO in regulating BMI is mechanistically unclear and has been called into question by studies suggesting linkage to variants in nearby genes IRX3 and IRX5 (Smemo et al. 2014; Claussnitzer et al. 2015). Loss-of-function (LoF) variants in the melanocortin 4 receptor (MC4R) are associated with obesity in humans (Krude et al. 1998; Vaisse et al. 1998; Yeo et al. 1998). Though a compelling target for body weight reduction, MC4R modulators are associated with adverse cardiovascular side effects (Yeo et al. 2021). Identification of additional BMI-associated genes may provide greater insight into the biology of body weight control and yield genes for which therapeutic modulation is tractable.

G protein coupled receptors (GPCRs) are attractive drug targets that have been associated with numerous phenotypes in human genetics studies and in mouse models (Erlandson et al. 2018). Loss-of-function in GPR151, a member of this class of receptor, has been associated with decreased BMI (Emdin et al. 2018; Tanigawa et al. 2019; Akbari et al. 2021). Additionally, at GPR151, carriage of rare (alternative allele frequency [aaf] < 1%) putative loss-of-function (plof) variants and bioinformatically predicted damaging missense variants have also been associated with a decrease in BMI (Akbari et al. 2021).

To date, homozygous plof carriers (human knockouts) have not been reported in detail. To follow up on the published genetic association for GPR151, we use the Pakistan Genome Resource (PGR), which is the world’s largest biobank of human homozygous plof carriers (knockouts) identified through whole-exome sequencing of >80,000 participants. Here, we (i) identify homozygous carriers of GPR151 plof variants, including those specific to South Asia, (ii) confirm *in vitro* that these variants are loss-of-function, (iii) test if GPR151 knockouts are associated with BMI, T2D, or other metabolic traits, and (iv) characterize GPR151^-/-^ mice for body weight.

## Results and Discussion

### Association of GPR151 plof variants with BMI and cardiometabolic events

The PGR at the Center for Non-Communicable Diseases (CNCD) in Pakistan is a large biobank of highly consanguineous participants. A medical history and numerous clinical measurements including BMI, T2D status, and myocardial infarction (MI) status, are available for most participants. In the PGR, we identified 48 homozygous carriers of three independent plof variants in GPR151 with a cumulative allele frequency of 2.2% (Table S1). Variants included Arg95Ter, as well as Tyr99Ter and Phe175LeufsTer7 that are both highly enriched in South Asia. By comparison, 21 homozygous plof carriers were identified among 281,852 exome-sequenced participants in the UK Biobank study and 13 homozygous plof carriers in non-south-Asian populations of gnomAD (Karczewski et al. 2020).

*GPR151* is expressed from a single exon, and nonsense substitutions are more likely to escape nonsense mediated decay (NMD) in single-exon genes. To determine if truncated proteins are expressed from GPR151 variant transgenes, we transiently transfected HEK293 cells with cDNA expression constructs corresponding to each variant with an N-terminal HA-tag. From transfected cells, total cell lysates and isolated membrane extracts were generated and evaluated by western blot (Figure 1). Wild type (WT) GPR151 protein expressed at high levels and was detected in both the total lysate and in membrane extracts. In contrast, Arg95Ter and Tyr99Ter were not detectable in either extract. Although, the Phe175LeufsTer7 variant protein was detectable in the both the total cell and membrane lysates and migrated at a size consistent with the expected truncation, it expressed at significantly lower levels compare to the wild-type, likely reflecting impaired stability. Our *in vitro* observations indicate that the GPR151 nonsense variants in PGR are loss-of-function alleles.

**Figure 1.**
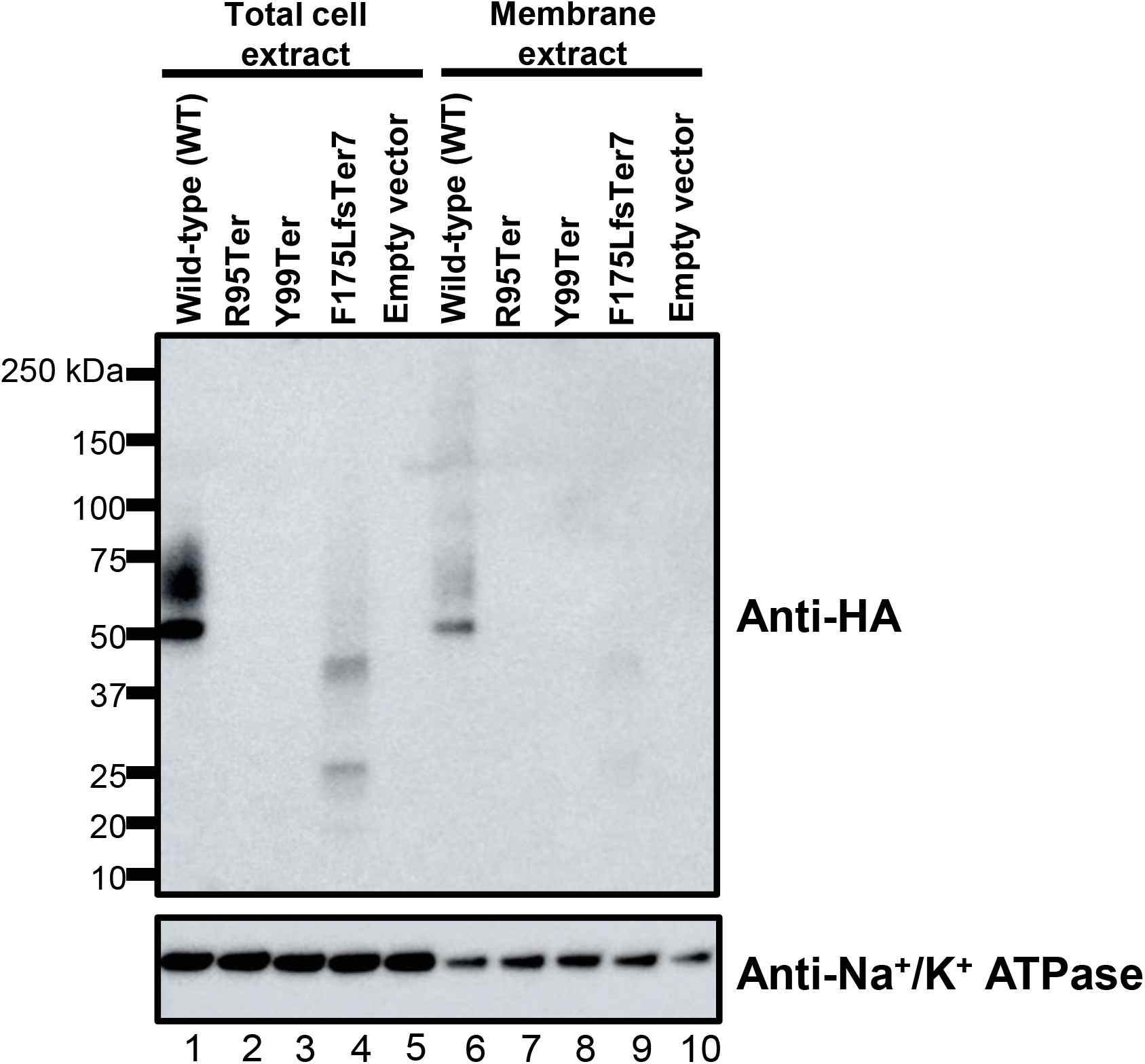
GPR151 variant proteins are not stably expressed. Western blot expression of HEK293 cells transfected with pcDNA3.1 plasmids encoding GPR151 variants with N-terminal HA-tag. Na^+^/K^+^ ATPase is shown as a loading control.

To test for associations with BMI, we analyzed plof variants individually and in a gene burden test. For our gene-burden analyses (caf = 2.2%), our study was adequately powered (80% at an α = 0.05) to detect a mean difference of 0.36 kg/m^2^ of BMI in knockouts compared to non-carriers. Similarly, for Tyr99Ter (aaf = 1.8%), our study was adequately powered (80% at an α = 0.05) to detect a mean difference of 0.40 units of BMI in knockouts compared to non-carriers. Neither the individual variants nor the plof gene burden showed statistically significant associations with BMI as compared to previous studies (Table 1). For the previously reported Arg95Ter and the South Asia-specific Tyr99Ter, knockouts were not found to have a significantly lower BMI compared to non-carriers. In human knockouts across all variants, we did not identify any clinically meaningful (>5%) reduction in BMI (26.7±4.4 kg/m^2^ in knockouts versus 26.3±3.3 kg/m^2^ in non-carriers). We tested for associations with other relevant traits including waist hip ratio, cholesterol and triglyceride levels: No significant associations or consistent trends were observed (Table S2).

**Table 1.** GPR151 associations with BMI.

| GRCh38 chr:pos | Reference allele | Alternate allele | HGVSp | Heterozygous carriers | Homozygous carriers | P | Beta [95% CI] kg/m<sup>2</sup> (additive) | Beta [95% CI] (knockouts only) |
| --- | --- | --- | --- | --- | --- | --- | --- | --- |
| 5:146515831 | G | A | Arg95Ter | 53 | 1 | 0.82 | -0.126<br>[-1.23 – 0.98] |  |
| 5:146515817 | G | T | Tyr99Ter | 945 | 34 | 0.92 | 0.0131<br>[-0.24 – 0.27] | -0.021<br>[-1.37 – 1.33] |
| 5:146515587 | CTA | C | Phe175LeufsTer7 | 92 | 3 | 0.28 | 0.406<br>[-0.32 – 1.14] |  |
| Gene Burden |  |  |  | 1,141 | 38 | 0.73 | 0.0405<br>[-0.20 – 0.28] | 0.431<br>[-0.99 – 1.85] |

*chr, chromosome; pos, position; HGVSp, Human Genome Variation Society protein level change; kg, kilograms; m, meter; CI, confidence interval*

Next, we analyzed the *GPR151* variants to assess reduction in T2D risk (Table 2). For gene-burden analyses, our study was powered to observe an odds ratio of T2D of 0.82 or lower (80% at an α = 0.05). Each plof variant was associated with a slight and non-significant increase in the risk for T2D. For the plof gene burden we observed a nominally significant increase in T2D (P=0.03, OR = 1.18 [1.02 – 1.37]). Similar analyses for MI risk did not yield any significant associations (Table S3).

**Table 2.**
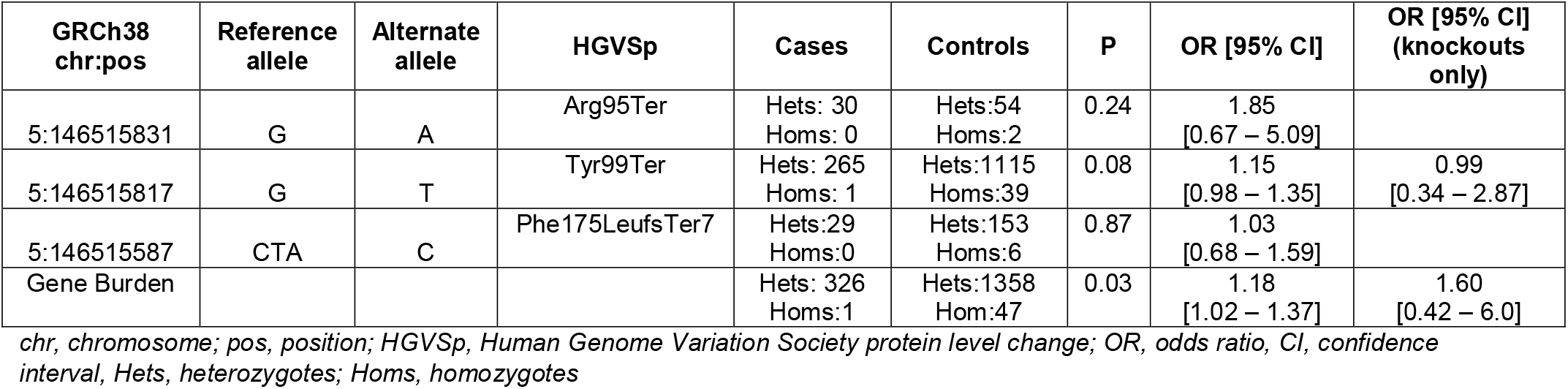
Association with T2D.

| GRCh38 chr:pos | Reference allele | Alternate allele | HGVSp | Cases | Controls | P | OR [95% CI] | OR [95% CI] (knockouts only) |
| --- | --- | --- | --- | --- | --- | --- | --- | --- |
| 5:146515831 | G | A | Arg95Ter | Hets: 30<br>Homs: 0 | Hets:54<br>Homs:2 | 0.24 | 1.85<br>[0.67 – 5.09] |  |
| 5:146515817 | G | T | Tyr99Ter | Hets: 265<br>Homs: 1 | Hets:1115<br>Homs:39 | 0.08 | 1.15<br>[0.98 – 1.35] | 0.99<br>[0.34 – 2.87] |
| 5:146515587 | CTA | C | Phe175LeufsTer7 | Hets:29<br>Homs:0 | Hets:153<br>Homs:6 | 0.87 | 1.03<br>[0.68 – 1.59] |  |
| Gene Burden |  |  |  | Hets: 326<br>Homs:1 | Hets:1358<br>Hom:47 | 0.03 | 1.18<br>[1.02 – 1.37] | 1.60<br>[0.42 – 6.0] |

*chr, chromosome; pos, position; HGVSp, Human Genome Variation Society protein level change; OR, odds ratio, CI, confidence interval, Hets, heterozygotes; Homs, homozygotes*

In total, complete loss of *GPR151* function in human knockouts was not associated with clinically meaningful changes in BMI or other traits related to obesity or metabolism, and was associated with a modest increase in T2D risk.

### Body weight in *Gpr151*^*-/-*^ mice

To determine if GPR151 plays a role in body weight regulation in mice, GPR151 knockout (*Gpr151*^*-/-*^) mice were generated using the CRISPR-Cas9 system. Loss of *Gpr151* expression was evaluated by *in situ* hybridization (ISH). In wild-type, *Gpr151*^*+/+*^ mice, *Gpr151* mRNA expression was primarily observed in the habenular nucleus in the brain, consistent with prior reports (Broms et al. 2015) and in mucosal cells of the ilium and jejunum (Figure 2A and 2B). *Gpr151* mRNA expression was not detected in *Gpr151*^*-/-*^ mice (Figure 2C and 2D).

**Figure 2.**
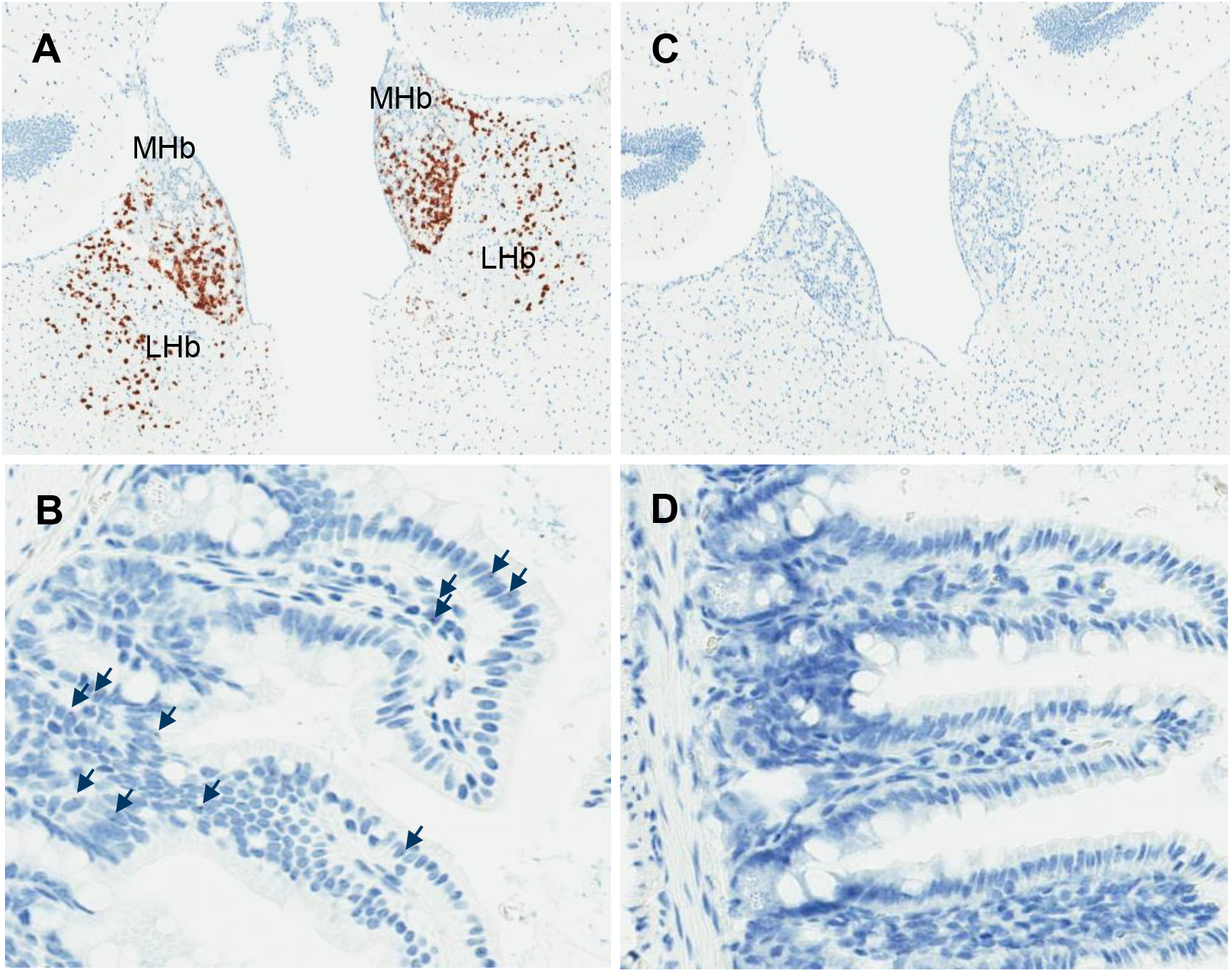
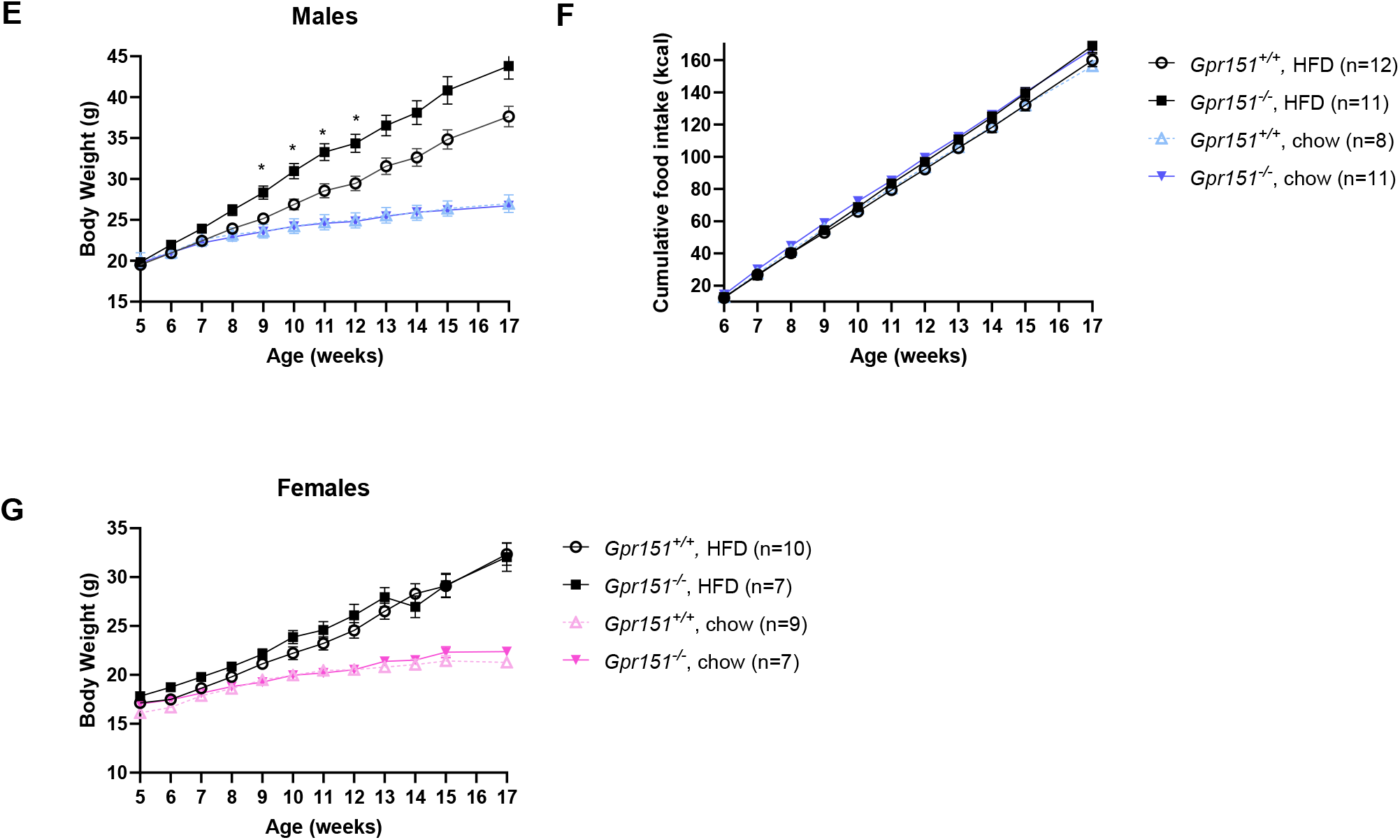
Male Gpr151^-/-^ mice gain weight on high fat diet. (A and C) Sections of mouse brain containing the medial habenula (MHb) and lateral habenula (LHb) from wild type and *Gpr151*^*-/-*^ mice, respectively, stained with a riboprobe for *Gpr151*. (B and D) Sections of mouse intestine containing the ileum and jejunum from wild type and *Gpr151*^*-/-*^ mice, respectively, stained with a riboprobe for *Gpr151*. Black arrows indicate cells containing *Gpr151* mRNA. (E) Body weights of male wild type and *Gpr151*^*-/-*^ mice on a standard chow diet and high fat diet. Data are presented as ± standard error of the mean (SEM). * p < 0.05, repeated measures, two-way ANOVA followed by post-hoc analysis using Sidak’s multiple comparisons test. (F) Cumulative food intake male wild type and *Gpr151*^*-/-*^ mice on a standard chow diet and high fat diet. Data are presented as ± SEM. (G) Body weights of female wild type and *Gpr151*^*-/-*^ mice on a standard chow diet and high fat diet. Data are presented as ± SEM.

Based on the published association between GPR151 plof and reduction in BMI, we predicted that the *Gpr151*^*-/-*^ mice would have lower body weight when compared to their wild type controls. On a standard chow diet, no difference in body weights was observed between *Gpr151*^*-/-*^ and wild type control mice of either sex (Figure 2E and 2G). *Gpr151*^*-/-*^ male mice fed a high fat diet for 12 weeks weighed ∼16% more than their wild type controls (Figure 2E). Food intake was similar between the two groups suggesting an alternative mechanism for diet-induced weight gain in the *Gpr151*^*-/-*^ male mice (Figure 2F). Female *Gpr151*^*-/-*^ mice fed a high fat diet gained the same amount of weight as their wild type counterparts suggesting a potential sex difference in the response to an obesogenic diet (Figure 2G).

In summary, complete loss of function of GPR151 is not associated with clinically meaningful change (i.e., > 5% change) in BMI. We identified homozygous carriers of the previously published Arg95Ter plof and additional South Asia-specific plof variants in PGR, confirmed that plof variants are unstable *in vitro*, and observed no statistically significant reduction in BMI in either heterozygous or homozygous carriers. Consistent with these observations, the body weights of male and female *Gpr151*^*-/-*^ mice were indistinguishable from wild type control mice on a standard chow diet, and were elevated in male *Gpr151*^*-/-*^ mice on a high fat diet without a corresponding increase in food intake. The preclinical model data indicate that the lack of association with BMI is generalizable rather than a human population-specific phenomenon. Our results also highlight the importance of taking into account the effect estimates and directionality of multiple loss of function variants when prioritizing GWAS results for functional follow-up. In aggregate, loss of GPR151 does not affect BMI in human knockouts in a clinically meaningful way and GPR151 antagonism is likely not a compelling therapeutic strategy for BMI reduction or T2D remission in humans.

## Materials and Methods

The Institutional Review Board (IRB) at the Center for Non-Communicable Diseases (IRB: 00007048, IORG0005843, FWAS00014490) approved the study. All participants gave informed consent.

### Variant QC and annotation

This study included a subset of 30,833 individuals with Whole Exome Sequencing and 9,292 individuals with Whole Genome Sequencing from the Pakistan Genome Resource. Samples with low allele balance for a variant (< 0.2) or low depth (< 10) were set to missing and variants that had a missingness rate > 5% were removed. We also removed variants failing VQSR filters or failing visual validation on IGV. Variants were annotated using Variant Effect Predictor (McLaren et al. 2016) based on the Ensembl101 gene model. We obtained a list of high-quality protein coding transcripts with annotated start and stop codons. Variants annotated as frameshift, stop gained, splice acceptor and splice donor variants are considered plof variants. Additionally, we filtered out plofs annotated as ‘low confidence’ according to the filtering criteria in LOFTEE (Karczewski et al. 2020). Cumulative Allele Frequency was calculated as described (Minikel et al. 2020).

### Case classification

Patients were categorized as T2D cases if they satisfied any one of the following criteria: 1) Documented history of diabetes, 2) HbA1c > 6.5%, 3) use of glucose lowering medication or 4) fasting glucose > 126 mg/dl. Patients were categorized as having had an MI as described previously (Saleheen et al. 2009).

### Statistical analysis

Associations with BMI (kg/m^2^), Cholesterol (mg/dl), triglycerides (mg/dl) and waist hip ratio were analyzed using multivariate linear regression adjusting for age, sex, age^2^ and top 5 genetic principal components (PCs). Associations with T2D and MI were analyzed using logistic regression, with Firth correction as implemented in glow (Mbatchou et al. 2021). Effect estimates from exomes and genomes data were meta analyzed using standard error weighted analysis as implemented in METAL (Willer et al. 2010). Power calculations were performed using Quanto v1.2 (Gauderman 2002).

### *In vitro* expression and western blot

Human codon optimized cDNAs corresponding to GPR151 wild-type (NCBI reference sequence NP_919227.2) and corresponding nonsense mutant constructs were cloned in pcDNA3.1(+) mammalian expression vectors with hemagglutinin (HA) epitope tags (YPYDVPDYA) appended to the amino termini. Transient transfection of adherent HEK293 cells was performed using Lipofectamine 2000 (Invitrogen) in a 6-well plate format according to manufacturer’s instructions. Cells were harvested 48-72 hours post-transfection by scraping, washed with PBS, and pelleted by centrifugation at 300xg for 5 min. Whole-cell lysates were prepared from half of each sample by SDS extraction. Cells were resuspended in PBS containing 2.5% (w/v) SDS, samples were incubated at 4°C for 10 minutes with end-over-end rotation, and insoluble material was removed by centrifugation at 16,000xg for 15 minutes. Membrane fractions were isolated from remaining cell sample using the Mem-PER Plus Membrane Protein Extraction Kit (Thermo Scientific) according to manufacturer’s instructions. Whole-cell lysates and isolated membrane fractions were analyzed by SDS PAGE and Western Blot against the HA epitope to detect GPR151 expression. The following antibodies were used: anti-HA monoclonal antibody (Clone 2-2.2.14, Invitrogen 26183); anti-Na+/K+ ATPase alpha-1 antibody (clone C464.6, Sigma 05-369); and anti-mouse IgG HRP-conjugated antibody (R&D Systems HAF007).

### Generation and phenotyping of *Gpr151*^*-/-*^ mice

Mice lacking the *Gpr151* gene were generated using the CRISPR-Cas9 system. All animal protocols were reviewed and approved by the Novartis Institutional Animal Care and Use Committee. The entire coding sequence of *Gpr151* is contained on a single exon (Ensembl gene ID# ENSMUSG00000042816). Two single guide RNA (sgRNA) sequences targeting sites just upstream of the translation start codon in exon 1 (ATCAAGCTCCTCCCTGCAGA) and within the 3’ untranslated region (3’ UTR) (TCATCAATATTGCTAAGCAG) were synthesized as crRNAs for Alt-R CRISPR-Cas9 system (Integrated DNA Technologies, Coralville, IA). A ribonucleoprotein mixture of the two crRNAs complexed with tracrRNA (Integrated DNA Technologies) and Cas9 protein (PNA Bio Inc, Newbury Park, CA) was electroporated into fertilized C57BL/6J embryos. The embryos were then implanted into pseudopregnant recipients. DNA lysates were prepared from tail biopsies of F0 generation pups using KAPA Mouse Genotyping Kit according to the manufacturer’s instructions (Kapa BioSystems, Cat# KK7302). Mice were genotyped by PCR using the following primers: For1 (5’-ACTTACAGACACTGTGAACAGC-3’) anneals to sequence upstream of Gpr151 exon 1, For2 (5’-TGGCTCCCAGAGTGGATAGC-3’) anneals to sequence within exon 1, and Rev1 (5’-TGCCTTTCTACTTACCAGGTTC-3’) anneals to sequence downstream of the Cas9 cut site within the 3’ UTR. For2 and Rev1 amplify a product of 614 bp corresponding to the wild-type allele, and For1 and Rev1 amplify a product of ∼233 bp corresponding to the null allele. PCR conditions were as follows: denaturation at 95°C for 3 min, 35 cycles of 15 sec at 95°C, 15 sec at 60°C, and 30 sec at 72°C, then 5 min at 72°C. F0 founders were bred to C57BL/6J mice for germline transmission of mutant alleles. The null allele of the F1 founder line selected was confirmed by Sanger sequencing (GeneWiz, South Plainfield, NJ) to have a deletion of 1343 bp between the expected Cas9 cleavage sites. Heterozygous mice were interbred to generate homozygous offspring for studies.

At six-weeks of age, *Gpr151*^*-/-*^ mice and wild type littermates were individually housed for body weight and food consumption measurements and provided either a standard chow diet (Purina Picolab 5053) or high fat diet in which 60% kcal is derived from fat (Research Diets D12492i) with *ad libitum* access to water. Female mice were grouped housed for body weight studies. Food intake and body weight were measured 1-2 times per week for 12 weeks following exposure to high fat diet. Animals were maintained on a 12-hour light/dark cycle. The study size is shown in Figure 2. One cohort was run for the study.

After 12 weeks of study, the mice were euthanized and brains collected to confirm GPR151 genotype by in situ hybridization. Samples from three animals of each genotype were used. Whole brains were fixed for 48 hours in 10% neutral buffered formalin and later embedded in paraffin. Four microns sections of each brain were collected. Staining was performed on the Leica Bond RX automated staining platform using the RNAscope 2.5 LSx Reagent Kit (ACDBio, Bio-Techne – 322440) for the following mouse *Gpr151* probe (ACDBio, Bio-Techne - 317328) following the standard RNAscope Assay (Wang et al. 2012).

## Data Availability

We will make summary level estimates relevant to this paper publicly available on our institutional website.

## Acknowledgements

We thank Jovita Marcinkeviciene, Ryan Streeper, Tim Kelly, and Igor Splawski for helpful discussions.

## Supplementary Tables

**Table S1.** List of all plof variants with homozygous carriers identified in 30,833 exomes and 9,292 genomes.

| GRCh38 chr:pos | Reference allele | Alternate allele | Allele frequency | Heterozygous carriers | Homozygous carriers | Variant effect | HGVSc | HGVSp |
| --- | --- | --- | --- | --- | --- | --- | --- | --- |
| 5:146515831 | G | A | 1.11E-03 | 85 | 2 | stop gained | c.283C>T | p.Arg95Ter |
| 5:146515817 | G | T | 1.84E-02 | 1394 | 40 | stop gained | c.297C>A | p.Tyr99Ter |
| 5:146515587 | CTA | C | 2.44E-03 | 184 | 6 | frameshift | c.525_526del | p.Phe175LeufsTer7 |

*chr, chromosome; pos, position; HGVSc and HGVSp, Human Genome Variation Society coding or protein level change, respectively*

**Table S2.** Association with waist-to-hip ratio and lipid related biomarkers.

| GRCh38 chr:pos | Reference allele | Alternate allele | Phenotype | HGVSp | Heterozygous carriers | Homozygous carriers | P | Beta [95% CI] (additive) |
| --- | --- | --- | --- | --- | --- | --- | --- | --- |
| plov Gene Burden |  |  | Cholesterol (mg / dl) |  | 1599 | 45 | 0.242 | 1.414 [-0.957 - 3.786] |
| 5:146515831 | G | A |  | Arg95Ter | 78 | 2 | 0.740 | 1.790 [-8.777 - 12.357] |
| 5:146515817 | G | T |  | Tyr99Ter | 1316 | 38 | 0.598 | 0.700 [-1.9 - 3.3] |
| 5:146515587 | CTA | C |  | Phe175LeufsTer7 | 171 | 5 | 0.383 | 3.161 [-3.94 - 10.26] |
| plov Gene Burden |  |  | Triglycerides (mg / dl) |  | 1591 | 45 | 0.925 | 0.274 [-5.384 - 5.93] |
| 5:146515831 | G | A |  | Arg95Ter | 79 | 2 | 0.581 | 7.060 [-18.014 - 32.133] |
| 5:146515817 | G | T |  | Tyr99Ter | 1307 | 38 | 0.855 | -0.580 [-6.784 - 5.623] |
| 5:146515587 | CTA | C |  | Phe175LeufsTer7 | 171 | 5 | 0.862 | -1.507 [-18.44 - 15.42] |
| plov Gene Burden |  |  | Waist Hip Ratio |  | 1263 | 42 | 0.567 | -0.001 [-0.004 - 0.002] |
| 5:146515831 | G | A |  | Arg95Ter | 56 | 2 | 0.147 | 0.010 [-0.004 - 0.024] |
| 5:146515817 | G | T |  | Tyr99Ter | 1035 | 37 | 0.222 | -0.002 [-0.005 - 0.001] |
| 5:146515587 | CTA | C |  | Phe175LeufsTer7 | 144 | 3 | 0.844 | -0.001 [-0.010 - 0.008] |

*chr, chromosome; pos, position; HGVSp, Human Genome Variation Society protein level change; kg, kilograms; m, meter; CI, confidence interval; mg/dl, milligrams per deciliter*

**Table S3.** Association with MI.

| GRCh38 chr:pos | Reference allele | Alternate allele | HGVSp | Cases | Controls | P | OR [95% CI] | OR [95% CI] (knockouts only) |
| --- | --- | --- | --- | --- | --- | --- | --- | --- |
| 5:146515831 | G | A | Arg95Ter | Hets: 50<br>Homs: 2 | Hets:35<br>Homs:0 | 0.33 | 1.39 [0.71 - 2.72] |  |
| 5:146515817 | G | T | Tyr99Ter | Hets: 736<br>Homs: 14 | Hets:658<br>Homs:26 | 0.90 | 0.99 [0.89 - 1.10] | 0.81 [0.38 - 1.71] |
| 5:146515587 | CTA | C | Phe175LeufsTer7 | Hets:100<br>Homs:0 | Hets:84<br>Homs:2 | 0.44 | 0.90 [0.68- 1.18] |  |
| Gene Burden |  |  |  | Hets: 913<br>Homs:20 | Hets:789<br>Hom:28 | 0.5 | 1.04 [0.39- 2.76] | 1.11 [0.5 - 2.4] |

*chr, chromosome; pos, position; HGVSp, Human Genome Variation Society protein level change; OR, odds ratio, CI, confidence interval*

